# Network state dynamics underpin craving in a transdiagnostic population

**DOI:** 10.1101/2023.10.03.23296454

**Authors:** Jean Ye, Kathleen A. Garrison, Cheryl Lacadie, Marc N. Potenza, Rajita Sinha, Elizabeth V. Goldfarb, Dustin Scheinost

**Author notes:** Send correspondence to Jean Ye, MRRC, 300 Cedar St, New Haven, CT 06519. These authors contributed equally (Elizabeth V. Goldfarb, Dustin Scheinost).

## Abstract

Emerging fMRI brain dynamic methods present a unique opportunity to capture how brain region interactions across time give rise to evolving affective and motivational states. As the unfolding experience and regulation of affective states affect psychopathology and well-being, it is important to elucidate their underlying time-varying brain responses. Here, we developed a novel framework to identify network states specific to an affective state of interest and examine how their instantaneous engagement contributed to its experience. This framework investigated network state dynamics underlying craving, a clinically meaningful and changeable state. In a transdiagnostic sample of healthy controls and individuals diagnosed with or at risk for craving-related disorders (N=252), we utilized connectome-based predictive modeling (CPM) to identify craving-predictive edges. An edge-centric timeseries approach was leveraged to quantify the instantaneous engagement of the craving-positive and craving-negative networks during independent scan runs. Individuals with higher craving persisted longer in a craving-positive network state while dwelling less in a craving-negative network state. We replicated the latter results externally in an independent group of healthy controls and individuals with alcohol use disorder exposed to different stimuli during the scan (N=173). The associations between craving and network state dynamics can still be consistently observed even when craving-predictive edges were instead identified in the replication dataset. These robust findings suggest that variations in craving-specific network state recruitment underpin individual differences in craving. Our framework additionally presents a new avenue to explore how the moment-to-moment engagement of behaviorally meaningful network states supports our changing affective experiences.

## Introduction

Communications between brain regions constantly flow to give rise to our changing experiences. Traditional neuroimaging analytic methods (e.g., static functional connectivity) tend to average data over several minutes, providing a static snapshot of brain-behavior associations. In contrast, brain dynamic approaches offer a glimpse into how brain responses fluctuate and interact to contribute to associated behaviors (1–3). This additional temporal information can be particularly beneficial when examining behaviors that evolve with time. Affective and motivational states serve as prominent examples (4–6). While these states wax and wane, they can also linger in some individuals (7). Persisting affective states can become maladaptive, preventing individuals from flexibly responding to ongoing demands. Regulating the duration of affective states is critical for well-being and psychopathology (8, 9). A comprehensive understanding of the temporal fluctuations in these emotional and motivational experiences requires similarly dynamic markers of brain responses. Such investigations can show how dynamic brain engagement supports these states and how this process may go awry in psychopathology (10).

One dynamic state with important implications for psychopathology is craving (11), which describes a strong desire, urge, or wanting. It is closely associated with maladaptive substance use and relapse (12–17). Like other affective/motivational states, craving ebbs and flows across time but can become a preoccupation in some individuals (18). This may result in perseverative, craving-dominant thoughts that are difficult to quell (19–21). The dynamic properties of craving indeed demonstrate important clinical relevance. In addition to craving intensity, dynamic changes in craving over time were linked to substance use (22–26). As time-varying changes in the brain support evolving affective/motivational experiences (5–6), there is a need to understand better how brain dynamics underlie craving.

Recent studies have identified brain activation (27) and functional connectivity patterns (28, 29) distinctively associated with craving. But little is known about how these brain regions dynamically interact across time to contribute to craving. For instance, a brain-based craving network significantly predicted craving in a transdiagnostic sample, highlighting contributions of the default mode, salience, and subcortical networks (28). However, since the employed approach required calculating functional connectivity over a minutes-long period, how the craving network’s temporal fluctuations give rise to this affective state remains elusive.

To address this gap, we present a new approach combining two cutting-edge methods: connectome-based predictive modeling (CPM; 30) and edge timeseries (31, 32). This framework allows us to identify behaviorally meaningful network states and track them at the resolution of individual time points. We first employed CPM to identify a craving-predictive network. Edge timeseries then assessed the engagement of this craving network at every time point during a separate scan. Two network states were delineated: one characterized by predominantly more coactivation in edges positively predicting craving, the other more coactivation in edges negatively predicting craving. We investigated the relationship between network state engagement and craving in a transdiagnostic sample of healthy controls (HCs) and individuals at risk for or diagnosed with craving-related conditions. The robustness of these associations were further tested by external validation in an independent group of HCs and individuals with alcohol use disorder (AUD). As craving is linked to difficulty in disengaging from craving-related thought content (19), we hypothesized that individuals with an elevated level of craving would show a higher propensity for persisting in a network state positively associated with craving while spending less time in a network state negatively associated with craving.

## Materials and methods

### Participants

Two independent functional magnetic resonance imaging (fMRI) datasets were analyzed here. Participants in both datasets first completed a baseline condition (no sensory input) followed by a task condition where a neutral/relaxed state was induced via distinct sensory modalities (**Supplementary Materials**). Participants listened to a personalized audio script in the imagery dataset to induce relaxation (28). For the *visual stimuli* dataset, individuals viewed a series of neutral/relaxing images (33, 34). We focused on the neutral condition in this study. Functional connectivity under this condition contributed the most to predicting individual differences in craving (28). Furthermore, basal craving was strongly associated with substance use (e.g., days of alcohol use) during relapse (17, 35).

Craving levels were assessed in both datasets, with key differences in sample and timing. In the *imagery* dataset, participants rated their craving or wanting of substance (i.e., alcohol or cocaine for individuals with alcohol or cocaine use disorder, respectively) or food at the moment (0 = “not at all” to 10 = “more than ever”). Ratings were obtained once each after baseline and task conditions. In the *visual stimuli* dataset, participants indicated their levels of craving from 1 to 9 (1 = “not at all” to 9 = “very much so”) at 1-minute intervals during baseline and task. Craving ratings collected after baseline and task conditions were averaged separately. Baseline craving was used to identify craving-predictive edges, whereas task craving was examined in brain dynamic analysis. The imagery dataset included a diverse range of participants, including healthy controls (HCs) and individuals with alcohol use disorder (AUD), cocaine use disorder (CUD), prenatal cocaine exposure, or obesity. The *visual stimuli* dataset recruited HCs and individuals with AUD.

### FMRI data preprocessing

Acquisition and imaging parameters for the two datasets have been detailed elsewhere (28, 33, 34). Standard preprocessing procedures described previously were applied to both datasets’ structural and functional data (see **Supplementary Materials** for more details). Parcellation was performed using the Shen-368 atlas plus 9 additional subcortical and brainstem structures (36). We excluded participants if any of their runs showed a mean framewise displacement over 0.2mm. Additional quality control criteria are summarized in the Supplementary Material. After exclusions, 252 *imagery* participants (88 female; Age:27.7±9.904) and 173 *visual stimuli* participants (79 female; Age: 31.902±10.867) were included in our analysis.

### Craving-predictive edges

To identify craving-predictive edges, we used mean baseline connectomes from the *imagery* dataset as input to CPM to predict craving collected immediately following the baseline scan (**Supplementary Materials**). This model was internally cross-validated using a leave-one-out design. In brief, linear regression was trained to predict craving with static functional connectome in all but one participant. The model was later applied to the leave-out participant to predict craving. Model performance was evaluated by correlating self-reported craving with predicted craving. Non-parametric p-values were computed using permutation testing. We repeated these steps after randomly shuffling craving measures across participants 500 times. Given that the *imagery* dataset included a larger and more diverse sample regarding craving-related disorders, it was selected to identify a craving network for our primary analysis (**Figure 1B**). CPM identified two networks: a positive subnetwork (i.e., stronger connectivity predicted higher craving) and a negative subnetwork (i.e., stronger connectivity predicted lower craving).

**Figure 1.**
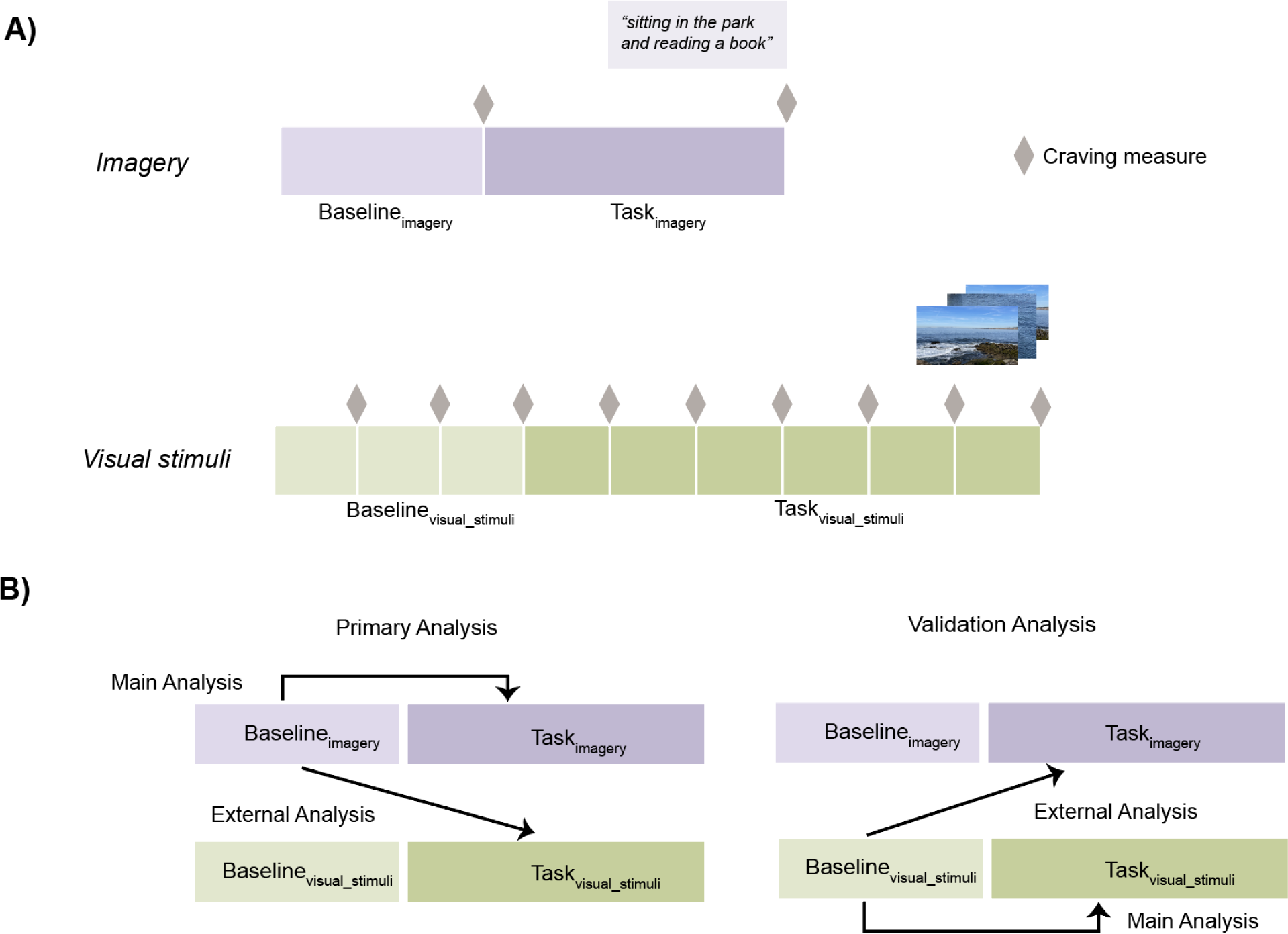
Datasets and analysis schematic. **A)** illustrates the two datasets analyzed in this study (*imagery* dataset in purple and *visual stimuli* dataset in green). Self-reported craving was collected at the end of each baseline or task run, as indicated by the gray diamonds. **B)** represents the analysis pipeline. Craving network was identified using the baseline condition of either the *imagery* or the *visual stimuli* dataset. Network state dynamics were examined in the task condition. For both primary and validation analysis, we first investigated brain dynamics in the task condition from the same dataset (main analysis) before extending the network to an independent dataset (external analysis).

### Network state dynamics: primary analysis

We next capitalized on a new approach known as edge timeseries (31, 32) to track the two craving subnetworks’ moment-to-moment engagement during task condition. In brief, edge timeseries describes the element-wise product of the activation timeseries from a pair of brain nodes after z-scoring (31, 32). Values are higher when the two nodes co-activate and lower when they activate in different directions. For each subnetwork, we computed edge timeseries for each edge before summing them to create a subnetwork timeseries (**Figure 2A**). We next subtracted the negative subnetwork timeseries from the positive one (**Figure 2A**). This state timeseries allowed us to identify the dominant network state at each time point by delineating two craving-specific network states: positive-network-dominant (PND; state timeseries > 0; **Figure 2B**) and negative-network dominant (NND; state timeseries < 0; **Figure 2B**). In other words, the PND state was characterized by more heightened coactivation in edges that positively predicted craving.

**Figure 2.**
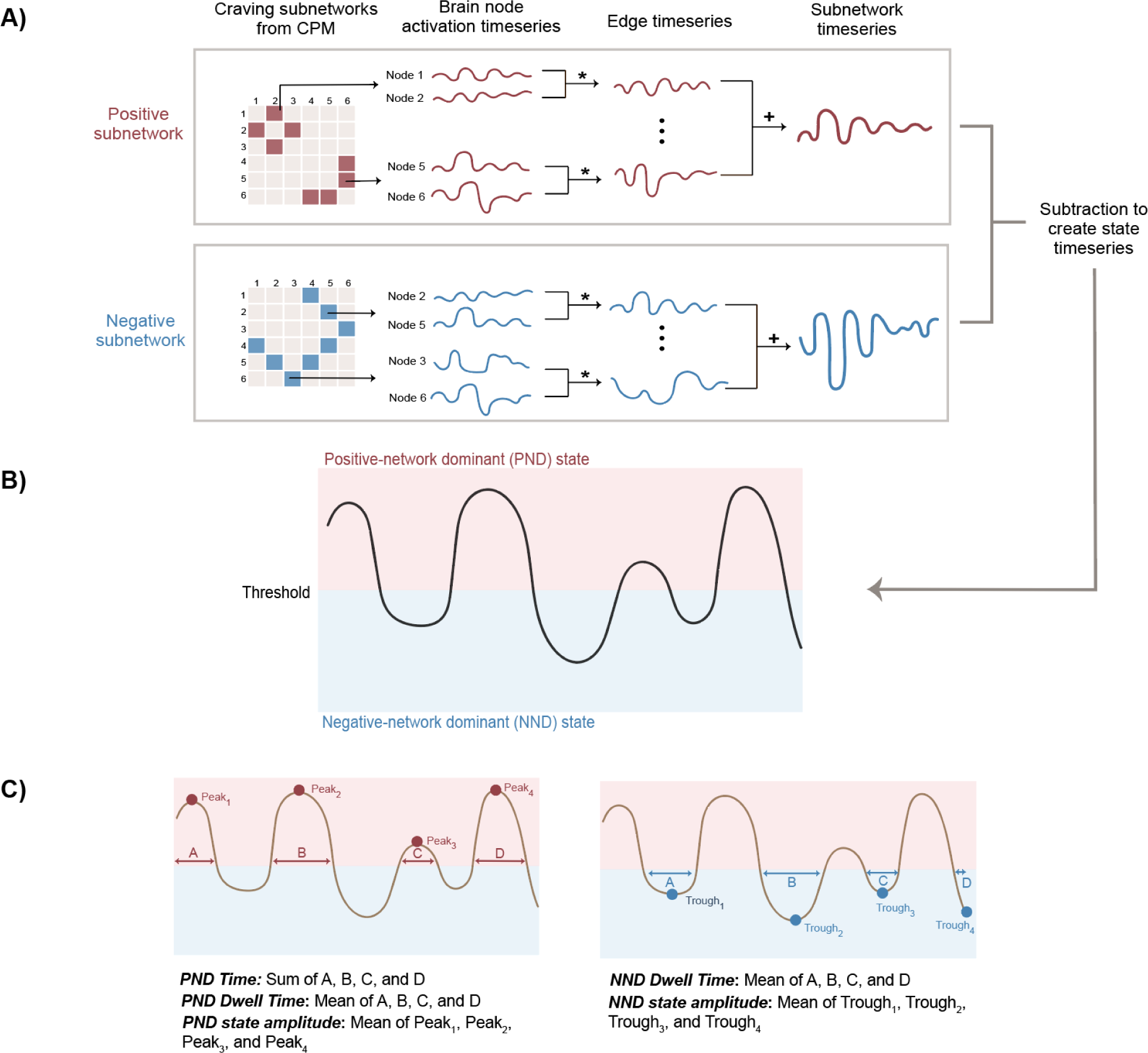
Brain dynamic measures. **A)** Methods overview. We first used CPM to identify a craving network and separated it into a positive and negative subnetwork. Edge timeseries tracked the moment-to-moment engagement of these two subnetworks. We additionally created a state timeseries by taking the difference between the positive and negative subnetworks. **B)** An example state timeseries (PND = red, NND = blue). **C**) Annotation of brain dynamic measures extracted from this state timeseries, including PND time, PND dwell time, PND state amplitude, NND dwell time, and NND state amplitude.

Using the state timeseries, the following network state dynamic measures can then extracted: 1) overall time in the PND state (i.e., the total number of time points where state timeseries was above 0); 2) dwell time for each state; and 3) state amplitude (i.e., peak and trough amplitude for the PND and NND state, respectively). To compute PND-related measures, we identified scan segments when an individual was in a PND state. Dwell time was operationalized as the number of consecutive time points spent in each segment. Peak amplitude was estimated by taking the absolute maximum value in the state timeseries within each segment. We then averaged these values to arrive at each participant’s mean dwell time and mean peak amplitude. NND-related measures were computed similarly. Instead of peak amplitude, the trough amplitude was extracted by finding the absolute value of the minimal value in the state timeseries. As amplitude describes the distance to the threshold separating the two states, we interpret a larger amplitude as a deeper network state. We hypothesized that individuals who reported a higher level of craving would spend more time in the PND state, dwell longer in the PND state, and express a deeper PND state. Simultaneously, they would dwell less in the NND state and show a shallower NND state (**Supplementary Figure 2** provides a visual representation).

To rigorously test these hypotheses, after defining craving subnetworks in the *imagery* baseline condition, we explored network state dynamics and their associations with craving in the *imagery* task condition using Pearson correlation (main analysis; **Figure 1B**). Following this main analysis, we externally validated the associations between craving and network state dynamics in an independent group of participants from the *visual stimuli* dataset (external analysis; **Figure 1B**). Specifically, the craving subnetworks identified in the *imagery* dataset were extended to the *visual stimuli* dataset. Task craving was next correlated with network state dynamics extracted from the *visual stimuli* task condition. FDR correction was performed on correlation analyses from main and external analyses.

### Network state dynamics: validation analysis

Finally, we switched the two datasets to test the robustness of our framework (validation analysis; **Figure 1B**). That is, the craving subnetworks were instead identified in the baseline condition of the *visual stimuli* dataset. Following the steps as described above, this new set of craving subnetworks was applied to the task condition of both the *visual stimuli* (main analysis) and *imagery* (external analysis) datasets to investigate the relationship between network and network state dynamics. We performed another FDR correction here as different subnetworks were utilized in our validation analysis. Similar results should appear in our main and validation analyses if our framework can reliably detect the relationship between network state dynamics and affective state.

## Results

### Primary main analysis: testing imagery craving network dynamics within the *imagery* dataset

We successfully predicted baseline craving using static connectivity in the *imagery* dataset (r=0.314; p=0.002; **Figure 3A & B; Supplementary Figure 4 & 5**). The positive and negative subnetworks contained 309 and 356 edges, respectively. Connections between the subcortical and visual systems and between the subcortical system and cerebellum were implicated in the positive subnetwork (**Figure 3B; Supplementary Figure 6**). The negative subnetwork included edges between the motor and visual systems and within visual systems (**Figure 3B; Supplementary Figure 6**).

**Figure 3.**
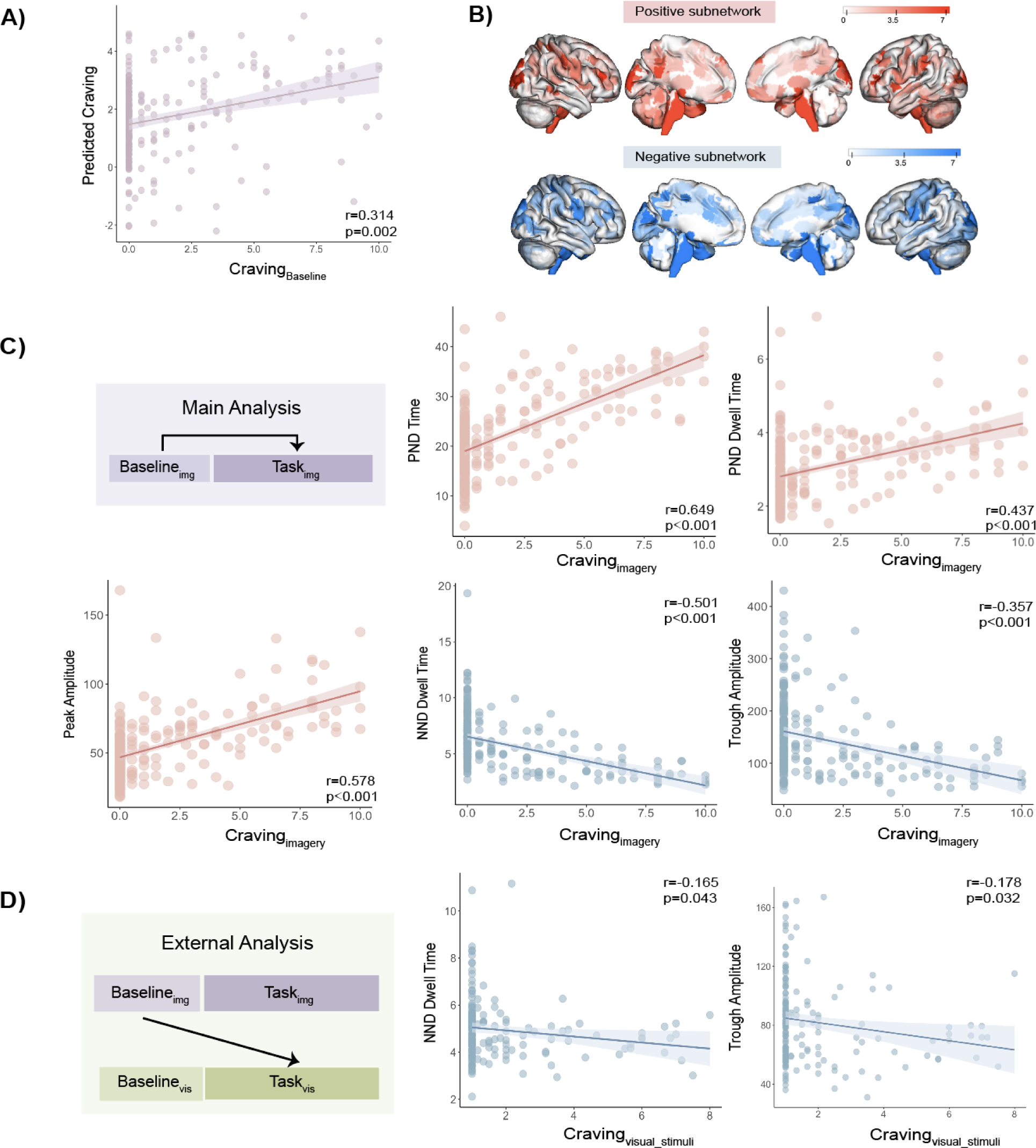
Primary Analysis. **A)** *Imagery* craving network predicted craving correlated significantly with self-reported craving. **B)** showed the node degrees (i.e., the number of edges each brain region contributed to the craving network) of the positive and negative subnetworks. **C)** During *imagery*, network state dynamics correlated with craving. Notably, some of the results were replicated during external validation in the *visual stimuli* dataset using the same imagery craving network **D)**. Baseline_img_, baseline condition from the *imagery* dataset; Task_img_, task condition from the *imagery* dataset; Baseline_vis_, baseline condition from the *visual stimuli* dataset; Task_vis_, task condition from the *visual stimuli* dataset.

Network state dynamic measures were extracted from the *imagery* task condition and correlated with post-task craving (**Supplementary Figure 4**; range: 0-10; mean±SD: 1.539±2.636). Individuals who reported higher craving spent more time in the PND state (r=0.649, p<0.001; **Figure 3C**), dwelled more in the PND state (r=0.437, p<0.001; **Figure 3C**), and demonstrated a deeper PND state (r=0.578, p<0.001; **Figure 3C**). Additionally, they dwelled less in the NND state (r=-0.501, p<0.001; **Figure 3C**) and showed a shallower NND state (r=-0.357, p<0.001; **Figure 3C**). These results indicate that elevated craving was linked to a sticky tendency to dwell in the PND state and a decreased propensity to occupy the NND state.

As a supplementary analysis, we next probed the specificity of the associations between craving and network state dynamics. First, with permutation testing, we assessed whether these patterns were specific to craving subnetworks or were also reflected in the general patterns of whole-brain network state dynamics (**Supplementary Materials**). Dynamics of the identified craving subnetwork demonstrated unique associations with craving (PND time: p=0.002; PND dwell: p=0.002; NND dwell: p=0.002; peak amplitude: p=0.002; trough amplitude: p=0.002). Another sensitivity analysis also revealed that none of the network state dynamic measures were correlated with heart rate, which was not significantly associated with craving during the neutral *imagery* condition (**Supplementary Table 1**).

### Primary external analysis: testing imagery craving network dynamics in the independent *visual stimuli* dataset

We next investigated whether the associations between network state dynamics and craving generalized to an external dataset. Network state dynamics were extracted during each *visual stimuli* task run. Mean dynamic measures across runs were subsequently correlated with mean craving during the task (range: 1-8; mean±SD: 1.854±1.635; **Supplementary Figure 4**). There were no significant associations between craving and any PND-state-related measures (PND time: r=0.090, p=0.301; PND dwell: r=-0.012, p=0.879; peak amplitude: r=-0.023, p=0.851). However, we successfully replicated our results with the NND state measures in this independent dataset. Individuals with higher craving dwelled less in the NND state (r=-0.165, p=0.043; **Figure 3D**) and demonstrated a shallower NND state (r=-0.178, p=0.032; **Figure 3D**). Furthermore, static functional connectivity of the two subnetworks defined in the imagery dataset was not significantly associated with mean craving in this new *visual stimuli* dataset (**Supplementary Materials**; positive: r=0.086, p=0.262; negative: r=-0.089, p=0.244; difference between subnetworks: r=0.100, p=0.190). These results suggest that network state dynamic measures may capture information about individual differences in craving that are not detectable using static functional connectivity measures alone.

### Validation main analysis: testing visual stimuli craving network dynamics within the *visual stimuli* dataset

As in the *imagery* dataset, we were able to identify a static craving network in the *visual stimuli* dataset that predicted baseline craving (r=0.256, p=0.004; **Figure 4A & B; Supplementary Figure 4 & 5**). The visual stimuli craving network contained substantially fewer edges than the imagery craving network (positive subnetwork: 175 edges; negative subnetwork: 142 edges; **Figure 4B**). Edges within the subcortical system, between the default mode network (DMN) and the salience network, and between the DMN and the frontoparietal network positively predicted craving (**Figure 4B; Supplementary Figure 7**). The negative subnetwork included edges within the DMN, between the cerebellum and salience network, and between the DMN and medial frontal network (**Figure 4B; Supplementary Figure 7**).

**Figure 4.**
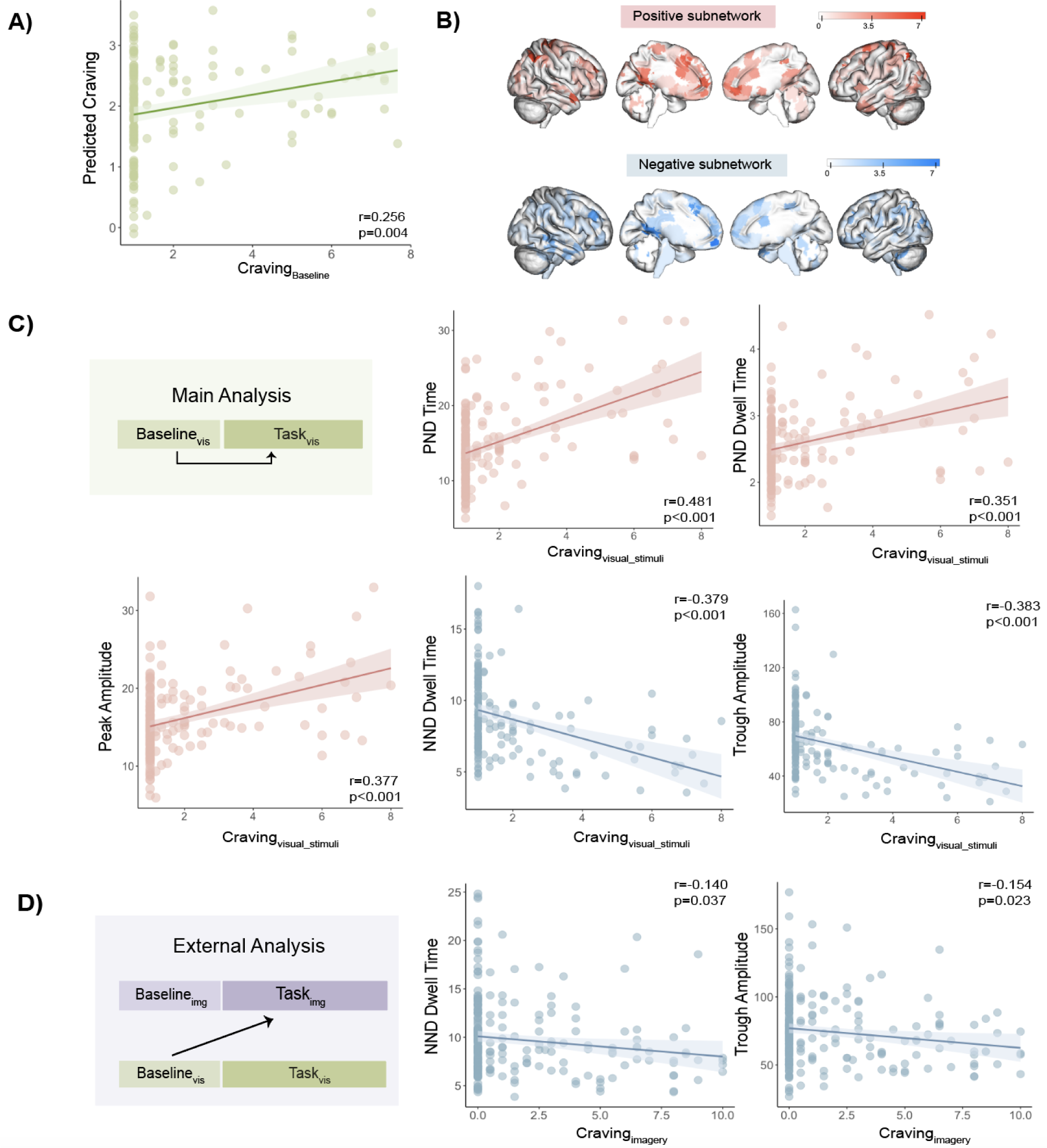
Validation analysis. **A)** Craving predicted by the visual stimuli craving network correlated significantly with self-reported craving. **B)** shows the node degree of the positive and negative sub-networks. **C)** During the task condition from the visual stimuli dataset, network state dynamics were associated with craving. When the visual stimuli craving network was extended to the imagery condition, we found that individuals with higher craving also dwelled more in the NND state and demonstrated a shallower NND state **D)**. Baseline_img_, baseline condition from the *imagery* dataset; Task_img_, task condition from the *imagery* dataset; Baseline_vis_, baseline condition from the *visual stimuli* dataset; Task_vis_, task condition from the *visual stimuli* dataset.

During the *visual stimuli* task condition, we found that individuals who reported higher craving spent more time in the PND state (r=0.481, p<0.001; **Figure 4C**), dwelled more in the PND state (r=0.351, p<0.001; **Figure 4C**), and showed a deeper PND state (r=0.377, p<0.001; **Figure 4C**). Additionally, they dwelled less in the NND state (r=-0.379, p<0.001; **Figure 4C**) and expressed a shallower NND state (r=-0.383, p<0.001; **Figure 4C**). Permutation testing again revealed that, compared to random edges in the brain, network state dynamics derived from the visual stimuli craving subnetworks offered unique insights into craving (PND time: p=0.002; PND dwell: p=0.002; NND dwell: p=0.002; peak amplitude: p=0.002; trough amplitude: p=0.002). We additionally investigated the associations between craving network dynamics and another behavioral measure not associated with craving, namely how focused participants were when performing the task (also rated on a scale from 1 to 9). Individual differences in focus were not significantly correlated with variations in network dynamic measures (**Supplementary Table 2**). These results further suggest that craving network dynamics conveyed specific information about our construct of interest.

### Validation external analysis: testing the visual stimuli craving network in the independent *imagery* dataset

We then extended the visual stimuli craving network to examine dynamics in the *imagery* task condition. Notably, despite the smaller visual stimuli craving networks, we replicated the NND state results. Individuals who reported a higher level of craving dwelled less in the NND state (r=-0.140, p=0.037) and expressed a shallower NND state (r=-0.154, p=0.023; **Figure 4D**). Craving was not significantly associated with PND-state-related dynamic measures (PND time: r=0.088, p=0.203; PND dwell: r=-0.054, p=0.438; peak: r=0.043, p=0.494). Unlike the primary analysis, static functional connectivity (based on the visual stimuli subnetworks) quantified during the *imagery* task condition was significantly associated with craving (positive: r=0.154, p=0.014; negative: r=-0.140, p=0.027; difference: r=0.166, p=0.008). Together with results from the primary analysis, these findings indicate that static functional connectivity and network state dynamic measures can provide complementary information about individual differences in background craving.

### Post-hoc investigations within craving cohorts

When we separated our participants into cohorts (i.e., HCs and individuals at risk for or diagnosed with craving-related conditions), we found that, as anticipated, basal craving differed significantly (**Supplementary Materials; Supplementary Figure 4**). Thus, we performed post-hoc analyses to investigate whether the relationships between craving and brain dynamics still held within each cohort. For our main analyses, craving remained significantly correlated with all brain dynamic measures within each cohort (**Supplementary Materials**). For our validation analyses, craving was only associated with brain dynamics when both cohorts were analyzed together (**Supplementary Materials**), likely due to increased statistical power from a larger sample size. Overall, the associations between brain dynamics and craving appear to be transdiagnostic.

## Discussion

We introduced a new framework to investigate how dynamic brain network state engagement underpinned the subjective experience of basal craving during neutral relaxing provocation in over 400 participants. Individuals with higher craving demonstrated sticky engagement of a network state positively associated with craving. Simultaneously, they dwelled less in a network state linked negatively to craving. These patterns were consistently observed in two independent datasets using different task paradigms. Network dynamics extracted from craving-specific edges contained unique information about craving but not other behavioral measures we examined. Our framework holds the potential to provide new insights into how the time-varying engagement of brain networks gives rise to the experience of affective and motivational states.

Our main goal was to examine relationships between network state dynamics and craving in a transdiagnostic sample. Thus, data from HCs and individuals at risk for or diagnosed with craving-related conditions were analyzed together. As results remained largely consistent when these phenotyped cohorts were examined separately, the observed dynamic brain/craving associations appeared to be transdiagnostic. Nevertheless, since greater craving and neural dysregulation have been consistently found in individuals at risk for or diagnosed with craving-related conditions (37–39), comparison of craving network dynamics using demographically matched cohorts remains an important direction for future research.

As anticipated, the relationships between network state dynamics and craving were more robust when craving subnetworks were identified within the same dataset. Notably, the links between craving and NND state dynamics were replicated in external validation. After identifying a brain network for which stronger connectivity predicted less craving, we found that individuals experiencing higher craving lingered less in a network state characterized by more heightened activation in this brain network. This pattern remained consistent regardless of the datasets used to identify craving subnetworks (i.e., *imagery* vs. *visual stimuli*) or the paradigm used to extract dynamic measures (i.e., naturalistic stimuli vs. traditional task designs). These findings suggest that compared to the PND state, the NND state may be more strongly associated with craving under the neutral condition explored here.

One plausible interpretation is that decreased NND state engagement reflects aberrant regulation of the connections between sensory processing and motor execution. The imagery negative subnetwork contained visual and visual/motor edges, whereas the visual stimuli negative subnetwork recruited salience/cerebellar edges. Differential connectivity between these regions has been observed in risky drinkers concerning stress (36). Our results were additionally consistent with past reports that people regularly consuming alcohol spent less time in a brain state made up of sensorimotor edges and demonstrated decreased dynamic functional connectivity between motor and visual regions (40).

Increased NND engagement might indicate better regulation between sensory processing and motor execution. This, in turns, may contribute to greater abilities or tendencies to inhibit automatic, habit-based behavioral responses linked to substance seeking (20). The interpretation that more sustained crosstalk between motor and sensory regions helps regulate automatic responding is supported by prior literature. Their increased recruitment positively predicted cocaine abstinence (41) and was associated with decreased impulsivity (42). Therefore, recruiting the NND state longer may permit more time for self-regulation to occur and interrupt potential undesirable impulsive or compulsive behaviors associated with craving (43, 44). Our interpretations suggest that individual differences in this self-regulation process may contribute to variations in basal craving, which has important implications for alcohol and substance misuse (45, 46). By shedding light on the neural mechanisms underpinning basal craving, the current investigations also reveal how aberrant inhibition and impulsive behavior may promote addictive engagement (44).

The network dynamic measures derived here are consistent with frameworks using concepts from nonlinear dynamical systems to investigate affective experiences (47, 48), psychopathology (49, 50), and substance use (51, 52). Following these studies, we may also consider the PND and NND states as attractors (i.e., stable states the brain can achieve). A shallower NND state (i.e., decreased dwell time and lower amplitude) was found in individuals experiencing higher craving. A shallower attractor can be more distractible (53). Echoing our interpretation earlier, this may be another indication that the NND state did not remain stable long enough to allow a careful evaluation of the triggered, more automatic sensory-motor pairing, thus resulting in higher craving. Individuals experiencing higher craving also expressed a deeper PND state (i.e., increased dwell time and larger amplitude). A deeper attractor is more stable, meaning that a system shows a higher propensity to stay in it and experiences greater difficulty transitioning out of it (49). The positive subnetworks from both datasets involved edges from the corticostriatal limbic circuitry. As these networks are important for craving (28, 54, 55), prolonged lingering in this network state matches a perseverative thought pattern occupied with craving-related content (21).

Altogether, individuals with higher craving simultaneously showed an excessively flexible NND state and an overly stable PND state. This imbalance between cognitive stability and flexibility might reflect impaired cognitive control, which has been closely linked to substance use across species (55, 56-58). The ability or tendency to efficiently balance stability and flexibility has been systematically examined using reversal learning paradigms, which probe the suppression of previously rewarded behavior and the adaptive shift to an alternative behavior in response to the changing environment (59). Impaired reversal learning has accompanied craving-related behaviors including cocaine use (60) and acted as a potential risk factor for future heavy drinking (61). Prenatal cocaine exposure has been associated with impaired reversal, with exposure to higher doses leading to worse performance (62). In line with these findings, networks important for cognitive control, including the frontoparietal network, the subcortical system, and the medial frontal network (59, 64, 65), were involved in the craving subnetworks identified in this study. This provides additional support for the possibility that individual variations in cognitive control may influence how stable or flexible network state engagement is within each participant.

Insights about how the dynamic engagement of network states related to craving are obscured from us without temporal information. Our framework extended prior work and provided a more comprehensive understanding of the neural mechanisms underlying craving. Edge timeseries is a recently introduced method that can track the moment-to-moment co-fluctuations of brain regions over time (31, 32). It offers several advantages over sliding-window, one of the most widely used dynamic functional connectivity methods. When using sliding-window, users need to specify window size, flexibility, and length (3). As functional connectivity is computed within each window, the window length must be long enough to limit spurious correlations and short enough to capture temporal information (65). Edge timeseries can provide edge engagement information at each time point without such specifications. This instantaneous tracking can be especially valuable when investigating evolving states such as craving.

The current study additionally extends previous research utilizing edge timeseries to identify network states to investigate individual differences (66). Instead of using brain activation level or canonical brain network atlases to characterize network states, we separated edges based on whether they positively or negatively predicted craving. The additional step of using CPM to select edges allowed us to identify network states specific to our behavior of interest and enhanced interpretability. Our framework retains the instantaneous temporal resolution from edge timeseries while providing interpretable, behaviorally relevant results given how the network states were identified with CPM. The consistent results reported here underscore our framework’s promise in capturing brain-and-behavioral information that complements prior methods. While we focused on craving, this framework can be readily applied to study how network state engagement supports other affective states (e.g., stress or anxiety) or trial-level task performance.It additionally offers flexibility to support combinations of varying tools. For instance, network-based statistics (67), instead of CPM, can be used to identify behaviorally relevant edges.

Several limitations of this study should be noted. As self-reported craving was only collected after each run or scan, investigating whether time-varying network engagement is associated with instantaneous craving remains an important topic for future study. Experimental work is needed to interpret our state amplitude measure. Do “deeper” state amplitude necessarily mean greater efforts in transitioning between network states? A craving regulation task may be leveraged to investigate whether an individual with a deeper PND state would also report greater difficulty disengaging. At the same time, our framework also presents new, exciting opportunities for future investigations. For instance, do peaks and troughs appear adjacent to important behavioral events (e.g., an exposure to craving-inducing stimuli or an attempt to regulate craving)? Studying how state engagement varies in response to such events may suggest future therapeutic targets by identifying key timepoints of dysregulated brain responses.

In summary, we developed a novel approach to investigate how spontaneous network state engagement is associated with the experience of an affective/motivational state with important clinical implications. In two independent datasets, increased basal craving was linked to an increased tendency to dwell in a craving-positive network state and a lower tendency to occupy a craving-negative network state. The framework introduced here presents a novel option to explore how time-varying changes in the brain support evolving affective or cognitive experiences.

## Acknowledgements

Supported by Yale Gruber Science Fellowship to J.Y.; Mind and Life Institute A-36649338 to K.A.G.; RL1-AA017539, R01 DA039136 to M.N.P.; R01-AA013892-15, P50-DA016556, UL1-DE019586, PL1-DA024859, R01-DK00039 to R.S.; K01-AA027832 to E.V.G; R01 MH121095 to D.S. We would like to thank Kelly Cosgrove, Yasmin Zakiniaeiz, and Ansel Hillmer for helpful discussions.

## Data availability

Data supporting findings of this study are available from the corresponding author upon reasonable request.

## References

1. Hutchison, R. M., Womelsdorf, T., Allen, E. A., Bandettini, P. A., Calhoun, V. D., Corbetta, M., Della Penna, S., Duyn, J. H., Glover, G. H., Gonzalez-Castillo, J., Handwerker, D. A., Keilholz, S., Kiviniemi, V., Leopold, D. A., de Pasquale, F., Sporns, O., Walter, M., & Chang, C. (2013). Dynamic functional connectivity: Promise, issues, and interpretations. NeuroImage, 80, 360–378.

2. Preti, M. G., Bolton, T. A., & Van De Ville, D. (2017). The dynamic functional connectome: State-of-the-art and perspectives. NeuroImage, 160, 41–54.

3. Lurie, D. J., Kessler, D., Bassett, D. S., Betzel, R. F., Breakspear, M., Kheilholz, S., Kucyi, A., Liégeois, R., Lindquist, M. A., McIntosh, A. R., Poldrack, R. A., Shine, J. M., Thompson, W. H., Bielczyk, N. Z., Douw, L., Kraft, D., Miller, R. L., Muthuraman, M., Pasquini, L., … Calhoun, V. D. (2020). Questions and controversies in the study of time-varying functional connectivity in resting fMRI. Network Neuroscience, 4(1), 30–69.

4. Tobia, M. J., Hayashi, K., Ballard, G., Gotlib, I. H., & Waugh, C. E. (2017). Dynamic functional connectivity and individual differences in emotions during social stress. Human Brain Mapping, 38(12), 6185–6205.

5. Gaviria, J., Rey, G., Bolton, T., Delgado, J., Van De Ville, D., & Vuilleumier, P. (2021). Brain functional connectivity dynamics at rest in the aftermath of affective and cognitive challenges. Human Brain Mapping, 42(4), 1054–1069.

6. Gaviria, J., Rey, G., Bolton, T., Ville, D. V. D., & Vuilleumier, P. (2021). Dynamic functional brain networks underlying the temporal inertia of negative emotions. NeuroImage, 240, 118377.

7. Kuppens, P., & Verduyn, P. (2017). Emotion dynamics. Current Opinion in Psychology, 17, 22–26.

8. Davidson, R. J. (2000). Affective style, psychopathology, and resilience: Brain mechanisms and plasticity. American Psychologist, 55(11), 1196–1214.

9. Houben, M., Van Den Noortgate, W., & Kuppens, P. (2015). The relation between short-term emotion dynamics and psychological well-being: A meta-analysis. Psychological Bulletin, 141(4), 901–930.

10. Kragel, P. A., Hariri, A. R., & LaBar, K. S. (2022). The Temporal Dynamics of Spontaneous Emotional Brain States and Their Implications for Mental Health. Journal of Cognitive Neuroscience, 34(5), 715–728.

11. Shiffman, S. (2000). Comments on craving. Addiction, 95(8s2), 171–175.

12. Hartz, D. T., Frederick-Osborne, S. L., & Galloway, G. P. (2001). Craving predicts use during treatment for methamphetamine dependence: A prospective, repeated-measures, within-subject analysis. Drug and Alcohol Dependence, 63(3), 269–276.

13. Weiss, R. D., Griffin, M. L., Mazurick, C., Berkman, B., Gastfriend, D. R., Frank, A., Barber, J. P., Blaine, J., Salloum, I., & Moras, K. (2003). The Relationship Between Cocaine Craving, Psychosocial Treatment, and Subsequent Cocaine Use. American Journal of Psychiatry, 160(7), 1320–1325.

14. Paliwal, P., Hyman, S. M., & Sinha, R. (2008). Craving predicts time to cocaine relapse: Further validation of the Now and Brief versions of the cocaine craving questionnaire. Drug and Alcohol Dependence, 93(3), 252–259.

15. Ferguson, S. G., & Shiffman, S. (2009). The relevance and treatment of cue-induced cravings in tobacco dependence. Journal of Substance Abuse Treatment, 36(3), 235–243.

16. Preston, K. L., Vahabzadeh, M., Schmittner, J., Lin, J.-L., Gorelick, D. A., & Epstein, D. H. (2009). Cocaine craving and use during daily life. Psychopharmacology, 207(2), 291–301.

17. Vafaie, N., & Kober, H. (2022). Association of Drug Cues and Craving With Drug Use and Relapse: A Systematic Review and Meta-analysis. JAMA Psychiatry, 79(7), 641–650.

18. Koob, G. F., & Volkow, N. D. (2016). Neurobiology of addiction: A neurocircuitry analysis. The Lancet Psychiatry, 3(8), 760–773.

19. Tiffany, S. T. (1999). Cognitive Concepts of Craving. Alcohol Research & Health, 23(3), 215–224.

20. Garavan, H., & Hester, R. (2007). The Role of Cognitive Control in Cocaine Dependence. Neuropsychology Review, 17(3), 337–345.

21. Tiffany, S. T., & Wray, J. M. (2012). The clinical significance of drug craving. Annals of the New York Academy of Sciences, 1248(1), 1–17.

22. Van Zundert, R. M., Ferguson, S. G., Shiffman, S., & Engels, R. (2012). Dynamic effects of craving and negative affect on adolescent smoking relapse. Health Psychology, 31(2), 226–234.

23. Preston, K. L., Kowalczyk, W. J., Phillips, K. A., Jobes, M. L., Vahabzadeh, M., Lin, J.-L., Mezghanni, M., & Epstein, D. H. (2018). Before and after: Craving, mood, and background stress in the hours surrounding drug use and stressful events in patients with opioid-use disorder. Psychopharmacology, 235(9), 2713–2723.

24. Motschman, C. A., Germeroth, L. J., & Tiffany, S. T. (2018). Momentary changes in craving predict smoking lapse behavior: A laboratory study. Psychopharmacology, 235(7), 2001–2012.

25. Ellis, J. D., Mun, C. J., Epstein, D. H., Phillips, K. A., Finan, P. H., & Preston, K. L. (2022). Intra-individual variability and stability of affect and craving among individuals receiving medication treatment for opioid use disorder. Neuropsychopharmacology, 47(10), Article 10.

26. Aslan, M., Sala, M., Gueorguieva, R., & Garrison, K. A. (2023). A Network Analysis of Cigarette Craving. Nicotine & Tobacco Research, 25(6), 1155–1163.

27. Koban, L., Wager, T. D., & Kober, H. (2023). A neuromarker for drug and food craving distinguishes drug users from non-users. Nature Neuroscience, 26(2), Article 2.

28. Garrison, K. A., Sinha, R., Potenza, M. N., Gao, S., Liang, Q., Lacadie, C., & Scheinost, D. (2023). Transdiagnostic Connectome-Based Prediction of Craving. American Journal of Psychiatry, 180(6), 445–453.

29. Antons, S., Yip, S. W., Lacadie, C. M., Dadashkarimi, J., Scheinost, D., Brand, M., & Potenza, M. N. (2023). Connectome-based prediction of craving in gambling disorder and cocaine use disorder. Dialogues in Clinical Neuroscience, 25(1), 33–42.

30. Shen, X., Finn, E. S., Scheinost, D., Rosenberg, M. D., Chun, M. M., Papademetris, X., & Constable, R. T. (2017). Using connectome-based predictive modeling to predict individual behavior from brain connectivity. Nature Protocols, 12(3), Article 3.

31. Faskowitz, J., Esfahlani, F. Z., Jo, Y., Sporns, O., & Betzel, R. F. (2020). Edge-centric functional network representations of human cerebral cortex reveal overlapping system-level architecture. Nature Neuroscience, 23(12), Article 12.

32. Zamani Esfahlani, F., Jo, Y., Faskowitz, J., Byrge, L., Kennedy, D. P., Sporns, O., & Betzel, R. F. (2020). High-amplitude cofluctuations in cortical activity drive functional connectivity. Proceedings of the National Academy of Sciences, 117(45), 28393–28401.

33. Sinha, R., Lacadie, C. M., Constable, R. T., & Seo, D. (2016). Dynamic neural activity during stress signals resilient coping. Proceedings of the National Academy of Sciences, 113(31), 8837–8842.

34. Blaine, S. K., Wemm, S., Fogelman, N., Lacadie, C., Seo, D., Scheinost, D., & Sinha, R. (2020). Association of Prefrontal-Striatal Functional Pathology With Alcohol Abstinence Days at Treatment Initiation and Heavy Drinking After Treatment Initiation. American Journal of Psychiatry, 177(11), 1048–1059.

35. Seo, D., Lacadie, C. M., Tuit, K., Hong, K.-I., Constable, R. T., & Sinha, R. (2013). Disrupted Ventromedial Prefrontal Function, Alcohol Craving, and Subsequent Relapse Risk. JAMA Psychiatry, 70(7), 727–739.

36. Goldfarb, E. V., Scheinost, D., Fogelman, N., Seo, D., & Sinha, R. (2022). High-Risk Drinkers Engage Distinct Stress-Predictive Brain Networks. Biological Psychiatry: Cognitive Neuroscience and Neuroimaging, 7(8), 805–813.

37. Sinha, R. (2013). The clinical neurobiology of drug craving. Current Opinion in Neurobiology, 23(4), 649–654.

38. Weissman, D. G., Schriber, R. A., Fassbender, C., Atherton, O., Krafft, C., Robins, R. W., Hastings, P. D., & Guyer, A. E. (2015). Earlier adolescent substance use onset predicts stronger connectivity between reward and cognitive control brain networks. Developmental Cognitive Neuroscience, 16, 121–129.

39. Bonson, K. R., Grant, S. J., Contoreggi, C. S., Links, J. M., Metcalfe, J., Weyl, H. L., Kurian, V., Ernst, M., & London, E. D. (2002). Neural Systems and Cue-Induced Cocaine Craving. Neuropsychopharmacology, 26(3), 376–386.

40. Vergara, V. M., Weiland, B. J., Hutchison, K. E., & Calhoun, V. D. (2018). The Impact of Combinations of Alcohol, Nicotine, and Cannabis on Dynamic Brain Connectivity. Neuropsychopharmacology, 43(4), Article 4.

41. Yip, S. W., Scheinost, D., Potenza, M. N., & Carroll, K. M. (2019). Connectome-Based Prediction of Cocaine Abstinence. American Journal of Psychiatry, 176(2), 156–164.

42. Herman, A. M., Critchley, H. D., & Duka, T. (2020). Trait Impulsivity Associated With Altered Resting-State Functional Connectivity Within the Somatomotor Network. Frontiers in Behavioral Neuroscience, 14.

43. Goldstein, R. Z., & Volkow, N. D. (2002). Drug Addiction and Its Underlying Neurobiological Basis: Neuroimaging Evidence for the Involvement of the Frontal Cortex. American Journal of Psychiatry, 159(10), 1642–1652.

44. Robbins, T. W., Gillan, C. M., Smith, D. G., Wit, S. de, & Ersche, K. D. (2012). Neurocognitive endophenotypes of impulsivity and compulsivity: Towards dimensional psychiatry. Trends in Cognitive Sciences, 16(1), 81–91.

45. Martins, J. S., Fogelman, N., Wemm, S., Hwang, S., & Sinha, R. (2022). Alcohol craving and withdrawal at treatment entry prospectively predict alcohol use outcomes during outpatient treatment. Drug and Alcohol Dependence, 231, 109253.

46. Wemm, S. E., Tennen, H., Sinha, R., & Seo, D. (2022). Daily stress predicts later drinking initiation via craving in heavier social drinkers: A prospective in-field daily diary study. Journal of Psychopathology and Clinical Science, 131(7), 780–792.

47. Scherer, K. R. (2009). Emotions are emergent processes: They require a dynamic computational architecture. Philosophical Transactions of the Royal Society B: Biological Sciences, 364(1535), 3459–3474.

48. Kuppens, P., Oravecz, Z., & Tuerlinckx, F. (2010). Feelings change: Accounting for individual differences in the temporal dynamics of affect. Journal of Personality and Social Psychology, 99(6), 1042–1060.

49. Rolls, E. T., Loh, M., & Deco, G. (2008). An attractor hypothesis of obsessive–compulsive disorder. European Journal of Neuroscience, 28(4), 782–793.

50. Rolls, E. T., Loh, M., Deco, G., & Winterer, G. (2008). Computational models of schizophrenia and dopamine modulation in the prefrontal cortex. Nature Reviews Neuroscience, 9(9), Article 9.

51. Epskamp, S., van der Maas, H. L. J., Peterson, R. E., van Loo, H. M., Aggen, S. H., & Kendler, K. S. (2022). Intermediate stable states in substance use. Addictive Behaviors, 129, 107252.

52. Kato, A., Shimomura, K., Ognibene, D., Parvaz, M. A., Berner, L. A., Morita, K., & Fiore, V. G. (2023). Computational models of behavioral addictions: State of the art and future directions. Addictive Behaviors, 140, 107595.

53. Loh, M., Rolls, E. T., & Deco, G. (2007). A Dynamical Systems Hypothesis of Schizophrenia. PLOS Computational Biology, 3(11), e228.

54. Sinha, R., Sinha, R., Li, C. S. R., Sinha, R., & Li, C. S. R. (2007). Imaging stress- and cue-induced drug and alcohol craving: Association with relapse and clinical implications. Drug and Alcohol Review, 26(1), 25–31.

55. Seo, D., & Sinha, R. (2014). Chapter 21—The neurobiology of alcohol craving and relapse. In E. V. Sullivan & A. Pfefferbaum (Eds.), Handbook of Clinical Neurology (Vol. 125, pp. 355–368). Elsevier.

56. Brockett, A. T., Pribut, H. J., Vázquez, D., & Roesch, M. R. (2018). The impact of drugs of abuse on executive function: Characterizing long-term changes in neural correlates following chronic drug exposure and withdrawal in rats. Learning & Memory, 25(9), 461–473.

57. Izquierdo, A., & Jentsch, J. D. (2012). Reversal learning as a measure of impulsive and compulsive behavior in addictions. Psychopharmacology, 219(2), 607–620.

58. Motzkin, J. C., Baskin-Sommers, A., Newman, J. P., Kiehl, K. A., & Koenigs, M. (2014). Neural correlates of substance abuse: Reduced functional connectivity between areas underlying reward and cognitive control. Human Brain Mapping, 35(9), 4282–4292.

59. Izquierdo, A., Brigman, J. L., Radke, A. K., Rudebeck, P. H., & Holmes, A. (2017). The neural basis of reversal learning: An updated perspective. Neuroscience, 345, 12–26.

60. Jentsch, J. D., Olausson, P., De La Garza, R., & Taylor, J. R. (2002). Impairments of Reversal Learning and Response Perseveration after Repeated, Intermittent Cocaine Administrations to Monkeys. Neuropsychopharmacology, 26(2), 183–190.

61. Shnitko, T. A., Gonzales, S. W., & Grant, K. A. (2019). Low Cognitive Flexibility as a Risk for Heavy Alcohol Drinking in Non-human Primates. *Alcohol (Fayetteville*, N.Y.), 74, 95–104.

62. Chelonis, J. J., Gillam, M. P., & Paule, M. G. (2003). The effects of prenatal cocaine exposure on reversal learning using a simple visual discrimination task in rhesus monkeys. Neurotoxicology and Teratology, 25(4), 437–446.

63. Cole, M. W., Repovš, G., & Anticevic, A. (2014). The Frontoparietal Control System: A Central Role in Mental Health. The Neuroscientist, 20(6), 652–664.

64. Ridderinkhof, K. R., Ullsperger, M., Crone, E. A., & Nieuwenhuis, S. (2004). The Role of the Medial Frontal Cortex in Cognitive Control. Science, 306(5695), 443–447.

65. Cohen, J. R. (2018). The behavioral and cognitive relevance of time-varying, dynamic changes in functional connectivity. NeuroImage, 180, 515–525.

66. Sporns, O., Faskowitz, J., Teixeira, A. S., Cutts, S. A., & Betzel, R. F. (2021). Dynamic expression of brain functional systems disclosed by fine-scale analysis of edge time series. Network Neuroscience, 5(2), 405–433.

67. Zalesky, A., Fornito, A., & Bullmore, E. T. (2010). Network-based statistic: Identifying differences in brain networks. NeuroImage, 53(4), 1197–1207.

