## Supplementary Materials for "Network state dynamics underpin craving in a transdiagnostic population"

Datasets

Two independent datasets were analyzed in this study and were described in detail in prior work (1, 2, 3). In the *imagery* dataset, participants began with a baseline condition where they were instructed to rest. This was followed by a task condition where they listened to a personalized script designed to induce relaxation and neutral feelings. Each run consisted of a baseline, an imagery, and a recovery condition. Two runs were collected. Data from the recovery condition were not analyzed here. The length of baseline condition varied between 43 to 62 volumes. All participants had 62 volumes in their imagery condition.

In the *visual stimuli* dataset, participants first completed three runs of baseline where they were presented with a flashing crosshair. This was followed by six task runs where individuals were instructed to passively view a series of neutral images. Each baseline or task run contained 60 volumes. Craving ratings were collected after each baseline and task run.

FMRI preprocessing

For both datasets, SPM12 was used to perform slice time and motion correction on the functional data. Additional data cleaning was completed using BioImage Suite. We regressed covariates of no interest, including linear and quadratic drift, mean white matter, cerebrospinal fluid (CSF), and gray matter signals, and a 24-parameter model of motion. The *visual stimuli* dataset included additional stick regressors for nonlinear motion outliers. Functional data were further temporally smoothed (cutoff frequency around ~0.12Hz). Timeseries data were parcellated using the Shen-368 atlas plus 9 additional subcortical and brainstem structures (36). The atlas was warped into single individual fMRI space using a series of linear and nonlinear registrations.

Quality control criteria

For the *imagery* dataset, we removed participants if they showed mean framewise displacement (MFD) over 0.2mm in either run, were missing self-reported craving data, or were missing timing information for us to separate the baseline and task conditions. After these exclusion criteria, 252 imagery participants were included (88 female; Age:27.7±9.904). This transdiagnostic sample consisted of healthy control participants (N=111), individuals with alcohol use disorder (AUD; N=35), individuals who reported binge drinking (N=8), individuals who reported heavy drinking (N=19), cocaine use disorder (CUD; N=28), prenatal cocaine exposure (N=35), and individuals with obesity or was overweight (N=16). We removed 21 brain nodes from this dataset due to missing coverage (**Supplementary Figure 1A**).

We removed participants for the visual stimuli dataset if they showed MFD over 0.2mm, did not have all nine imaging runs or craving ratings, or were missing volume in any of their runs. After these exclusion criteria, 173 participants were included (79 female; Age: 31.902±10.867), including 67 individuals with AUD and 106 healthy control participants. This dataset excluded two brain nodes due to missing coverage (**Supplementary Figure 1B**).

A)

B)

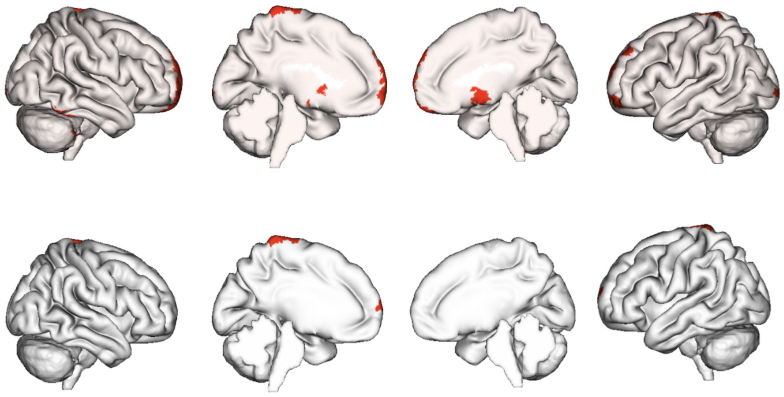

**Supplementary Figure 1**. Brain nodes excluded from the imagery **A)** and visual stimuli **B)** datasets due to missing coverage.

<u>Quality control after extracting network dynamic measures</u>

We excluded one *imagery* run from one participant and one *visual stimuli* run from six participants since they appeared to spend the entire run in the NND state. These runs were not analyzed due to concerns that participants may have gotten distracted or fallen asleep.

<u>Static functional connectivity analysis</u>

In our validation analysis, we also examined whether the craving network's static functional connectivity correlated with craving. We first computed static functional connectomes within each imagery or visual stimuli task condition run before averaging across runs. We then summed the strength of all edges in the positive or negative craving subnetwork before correlating the total network strength with craving across participants. We also took the difference between total positive and negative subnetwork strength to further match our dynamic analysis to assess its relationship with craving.

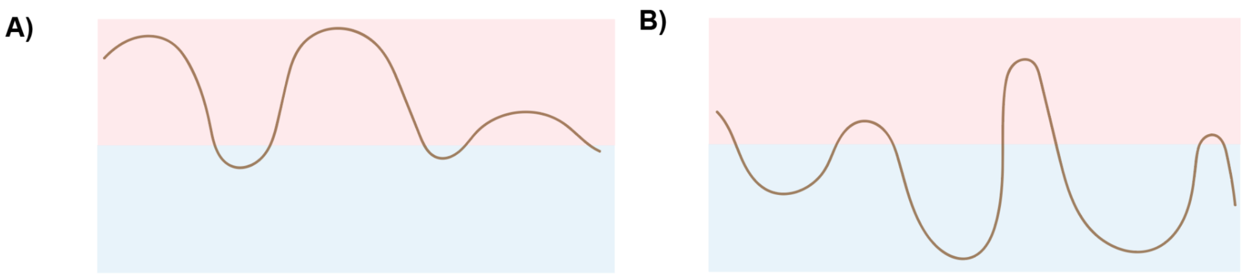


**Supplementary Figure 2. Cartoon examples of someone with a high A) or B) low level of craving.** A participant with more craving might show increased overall and dwell time in the PND state. They might also demonstrate a deeper PND state and a shallower NND state, indicating an increased difficulty in switching away from the PND state and maintaining the NND state. The opposite pattern might be observed in someone with a lower level of craving. They might dwell more in the NND state and show a deeper NND state but a shallower PND state.

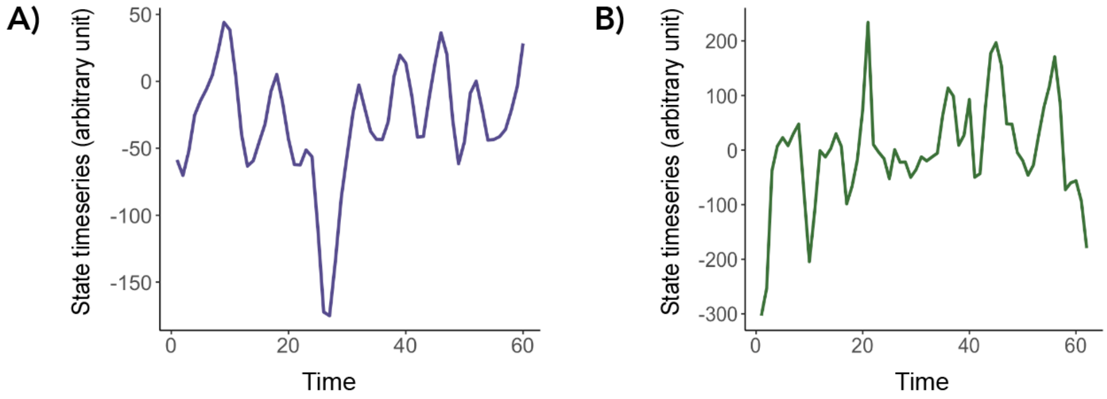


**Supplementary Figure 3**. Example state timeseries data from one imagery run **A)** and one visual stimuli run **B)**.

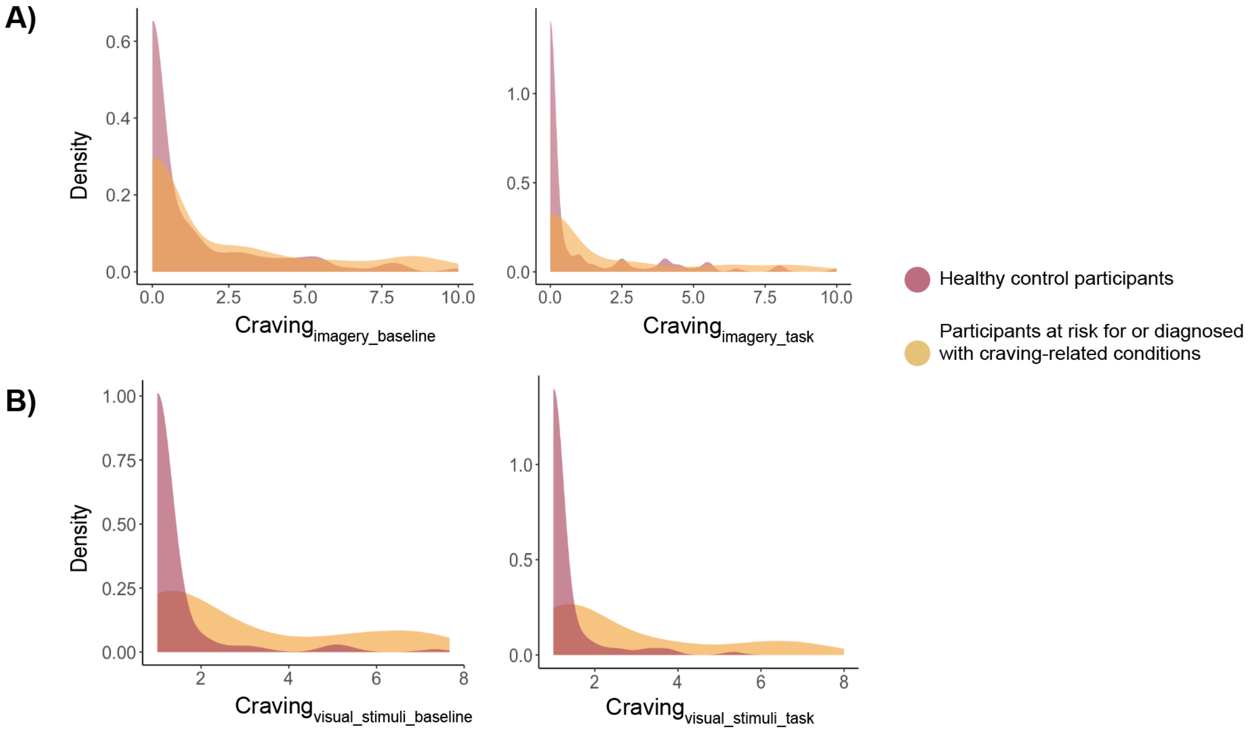


**Supplementary Figure 4. Density plot for craving measures.** Distributions of craving measures from the *imagery* and *visual stimuli* datasets are shown in **A)** and **B)**, respectively. Baseline craving was used to identify craving networks. The relationship between brain dynamics and craving was explored using craving measures collected after task condition.

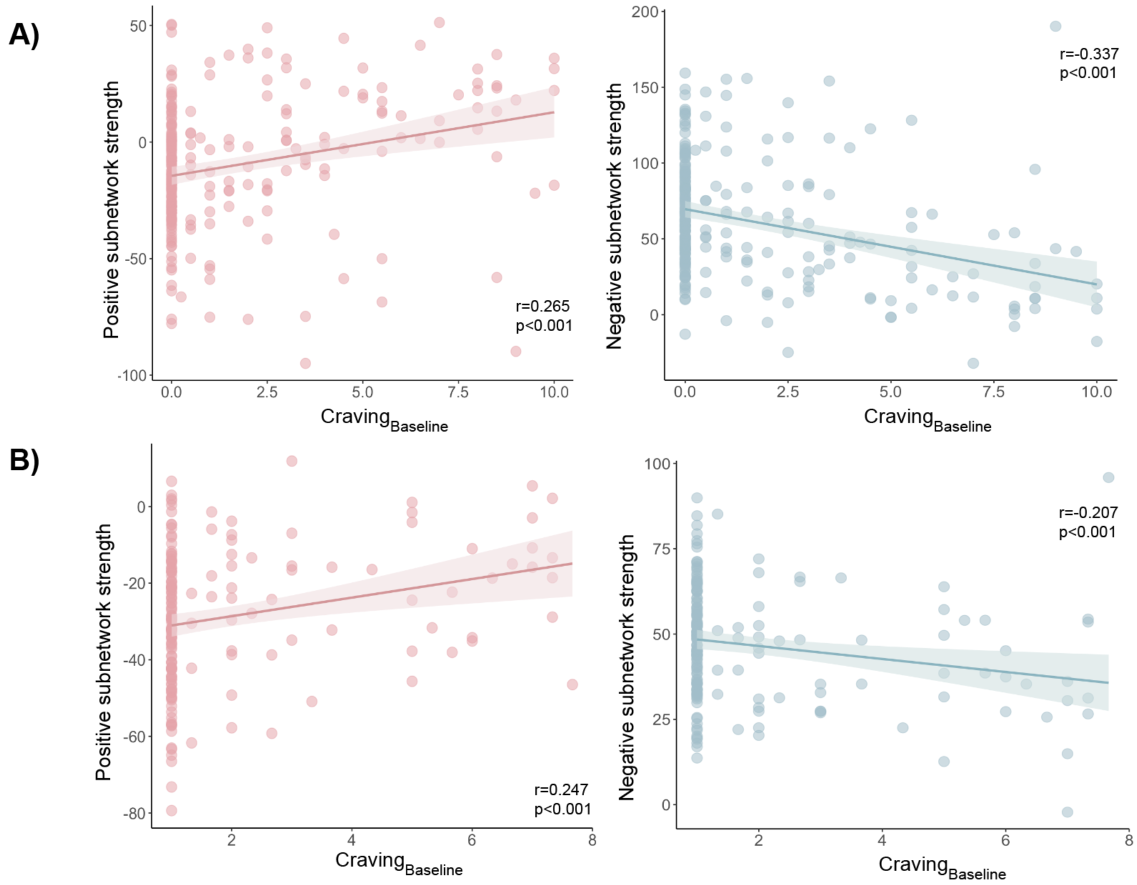


**Supplementary Figure 5.** Mean negative and positive subnetwork strength used to predict craving correlated with self-reported craving in the primary A) and validation analysis B).

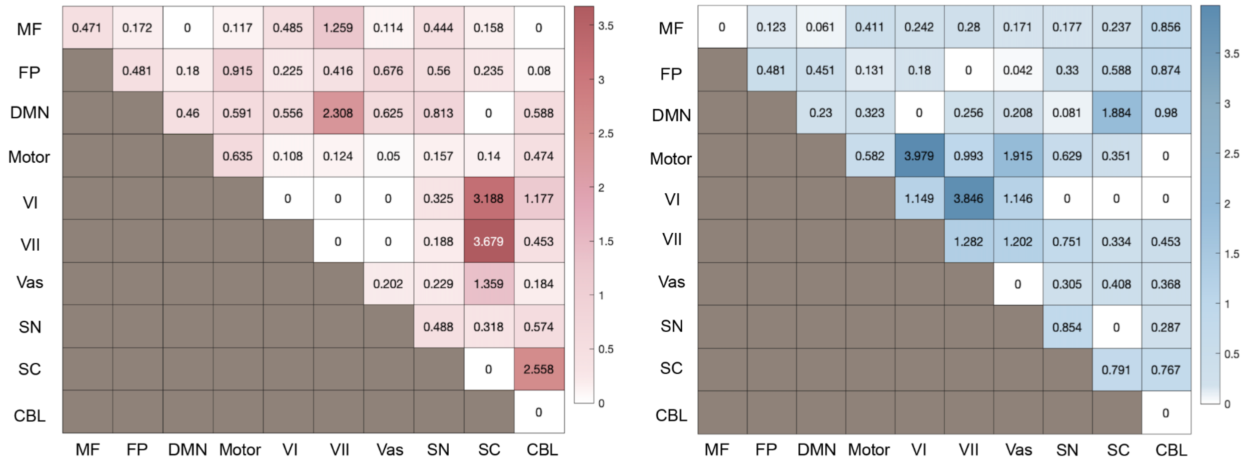


**Supplementary Figure 6**. Network edges involved in the imagery craving network. The number of significant edges were normalized (dividing the number of edges by the total number of possible edges within or between networks and timing by 100).

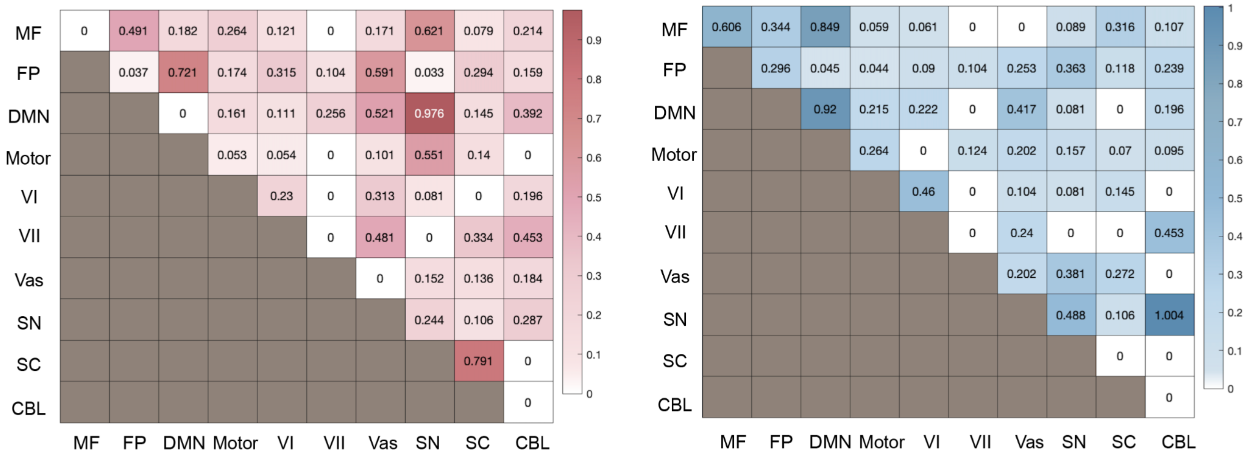


**Supplementary Figure 7**. Network edges involved in the visual stimuli craving network. The number of significant edges were normalized (dividing the number of edges by the total number of possible edges within or between networks and timing by 100).

**Supplementary Table 1.** Specificity analysis to examine whether craving-network dynamic measures were correlated with heart rate in the *imagery* dataset. Heart rate was not associated with craving (r=-0.033; p=0.613). Eight *imagery* participants did not have heart rate data and were excluded from this analysis.

| | Everyone (N=244) |
|---|---|
| PND time | r=-0.010; p=0.876 |
| PND dwell | r=0.047; p=0.462 |
| NND dwell | r=0.088; p=0.171 |
| Peak | r=-0.005; p=0.935 |
| Trough | r=0.023; p=0.726 |

**Supplementary Table 2.** Specificity analysis to examine whether craving-network dynamic measures were correlated with focus in the visual stimuli dataset. Focus was not associated with craving (r=0.140; p=0.067). Focus was analyzed in the visual stimuli dataset since heart rate was not available from this dataset. One *visual stimuli* participant did not have focus data and was excluded from this analysis.

| | Everyone (N=172) |
|---|---|
| PND time | r=-0.102; p=0.183 |
| PND dwell | r=-0.097; p=0.207 |
| NND dwell | r=0.105; p=0.170 |
| Peak | r=-0.073; p=0.340 |
| Trough | r=0.084; p=0.274 |

Exploring craving and network state dynamics within cohorts

We performed post-hoc analyses to examine whether the same associations between craving and network state dynamics can be consistently found within each cohort separately. Following prior work (1), we first separated the participants in the *imagery* dataset into healthy control participants (N=111) and individuals at risk for or diagnosed with craving-related conditions (N=141; 35 individuals with alcohol use disorder, 8 participants who reported binge drinking, 19 participants who reported heavy drinking, 28 participants with cocaine use disorder, 16 participants with obesity or was overweight, and 35 participants with prenatal cocaine exposure). We additionally separated the *visual stimuli* dataset into healthy control participants (N=106) and individuals with alcohol use disorder (N=67).

In both datasets, craving differed significantly between healthy controls and individuals at risk for or diagnosed with craving-related conditions (imagery: $t(250)=3.048$, $p=0.003$; visual stimuli: $t(171)=7.137$, $p<0.001$). Consistent patterns between craving and network state dynamics were observed within cohorts in the main analyses (**Supplementary Table 3 & 4**). But for external analysis (**Supplementary Table 5 & 6**), the associations between network state dynamics and craving were only significant when both cohorts were combined (see **Results**). These findings further emphasize the transdiagnostic nature of the association between craving and network state dynamics. While most correlations did not show significant cohort differences, the association between craving and trough amplitude differed significantly in the *visual stimuli* dataset (**Supplementary Table 4**). Future research should further investigate whether the relationship between craving and network state dynamics varies significantly in individuals with substance use disorder. As the participants included in the current study were not demographically matched and might have other conditions not considered here, we did not formally compare brain dynamics between cohorts in the current study.

**Supplementary Table 3. Primary main analysis within each group**

| | Individuals at risk for or diagnosed with craving-related conditions (N=141) | Healthy control participants (N=111) | Group interactions |
|---|---|---|---|
| PND time | r=0.677; p<0.001 | r=0.614; p<0.001 | z=0.84; p=0.401 |
| PND dwell | r=0.434; p<0.001 | r=0.487; p<0.001 | z=-0.52; p=0.603 |
| Peak amplitude | r=0.568; p<0.001 | r=0.637; p<0.001 | z=-0.84; p=0.401 |
| NND dwell | r=-0.528; p<0.001 | r=-0.448; p<0.001 | z=-0.82; p=0.412 |
| Trough amplitude | r=-0.381; p<0.001 | r=-0.309; p<0.001 | z=-0.64; p=0.522 |

**Supplementary Table 4. Validation main analysis within each group**

| | Individuals with alcohol use disorder (N=67) | Healthy control participants (N=106) | Group interactions |
|---|---|---|---|
| PND time | r=0.503; p<0.001 | r=0.320; p<0.001 | z=1.39; p=0.165 |
| PND dwell | r=0.369; p=0.002 | r=0.233; p=0.016 | z=0.94; p=0.347 |
| Peak amplitude | r=0.427; p<0.001 | r=0.220; p=0.023 | z=1.46; p=0.072 |
| NND dwell | r=-0.462; p<0.001 | r=-0.233; p=0.017 | z=-1.65; p=0.099 |
| Trough amplitude | r=-0.504; p<0.001 | r=-0.220; p=0.024 | z=-2.08; p=0.038 |

**Supplementary Table 5. Primary external analysis within each group**

| | Patients (N=67) | HCs (N=106) |
|---|---|---|
| PND time | r=0.095; p=0.444 | r=0.113; p=0.249 |
| PND dwell | r=0.032; p=0.799 | r=0.066; p=0.503 |
| Peak amplitude | r=0.019; p=0.881 | r=0.033; p=0.741 |
| NND dwell | r=-0.201; p=0.104 | r=-0.059; p=0.551 |
| Trough amplitude | r=-0.191; p=0.121 | r=-0.120; p=0.221 |

**Supplementary Table 6. Validation external analysis within each group**

| | Patients (N=141) | HCs (N=111) |
|---|---|---|
| PND time | r=0.116; p=0.172 | r=0.060; p=0.535 |
| PND dwell | r=-0.113; p=0.181 | r=0.088; p=0.357 |
| Peak amplitude | r=0.060; p=0.484 | r=0.023; p=0.808 |
| NND dwell | r=-0.161; p=0.056 | r=-0.122; p=0.203 |
| Trough amplitude | r=-0.163; p=0.053 | r=-0.117; p=0.222 |

**References**


1. Garrison, K. A., Sinha, R., Potenza, M. N., Gao, S., Liang, Q., Lacadie, C., & Scheinost, D. (2023). Transdiagnostic Connectome-Based Prediction of Craving. *American Journal of Psychiatry*, *180*(6), 445–453.
2. Sinha, R., Lacadie, C. M., Constable, R. T., & Seo, D. (2016). Dynamic neural activity during stress signals resilient coping. *Proceedings of the National Academy of Sciences*, *113*(31), 8837–8842.
3. Blaine, S. K., Wemm, S., Fogelman, N., Lacadie, C., Seo, D., Scheinost, D., & Sinha, R. (2020). Association of Prefrontal-Striatal Functional Pathology With Alcohol Abstinence Days at Treatment Initiation and Heavy Drinking After Treatment Initiation. *American Journal of Psychiatry*, *177*(11), 1048–1059.
